# Disinfection of N95 Respirators with Ozone

**DOI:** 10.1101/2020.05.28.20097402

**Authors:** Edward P. Manning, Matthew D. Stephens, Sannel Patel, Sylvie Dufresne, Bruce Silver, Patricia Gerbarg, Zach Gerbarg, Charles Dela Cruz, Lokesh Sharma

## Abstract

The coronavirus disease 2019 crisis is creating a shortage of personal protective equipment (PPE), most critically, N95 respirators for healthcare personnel. Our group was interested in the feasibility of ozone disinfection of N95 respirators as an alternative for healthcare professionals and organizations that might not have access to other disinfection devices. We tested the effectiveness of ozone on killing *Pseudomonas aeruginosa* (PsA) on three different N95 respirators: 3M 1860, 3M 1870, and 3M 8000. We used an ozone chamber that consisted of: an airtight chamber, an ozone generator, an ozone destruct unit, and an ozone UV analyzer. The chamber was capable of concentrating ozone up to 500 parts per million (ppm) from ambient air, creating an airtight seal, and precisely measuring ozone levels within the chamber. Exposure to ozone at 400 ppm with 80% humidity for two hours effectively killed bacteria on N95 respirators, types 1860, 1870, and 8000. There were no significant changes in filtration efficiency of the 1860 and 1870 type respirators for up to ten cycles of ozone exposure at similar conditions. There was no change in fit observed in the 1870 type respirator after ozone exposure. There was no significant change in the strap integrity of the 1870 type respirator after ozone exposure. Tests for filtration efficiency were not performed on the 8000 type respirator. Tests for fit or strap integrity were not performed on the 8000 or 1860 type respirators. This study demonstrates that an ozone application achieves a high level of disinfection against PsA, a vegetative bacteria that the CDC identifies as more difficult to kill than medium sized viruses such as SARS-CoV-2 (Covid-19). Furthermore, conditions shown to kill these bacteria did not damage or degrade respirator filtration. This is the first report of successful disinfection of N95 PPE with ozone of which the authors are aware. It is also the first report, to the authors’ knowledge, to identify necessary conditions for ozone to kill organisms on N95 masks without degrading the function of N95 filters.

## BACKGROUND

### Reuse of Personal Protective Equipment

The coronavirus disease 2019 (COVID-19) crisis created a shortage of personal protective equipment (PPE), most critically, NIOSH-certified N95 filtering facepiece respirators (commonly called “N95 respirators”) for healthcare personnel (HCP). “All FDA-cleared N95 respirators are labeled as ‘single-use’, disposable devices”(U.S. Food and Drug Administration, 2020h). Nevertheless, as supplies dwindled, front-line medical workers had no choice but to reuse respirators and experiment with “home recipes” for disinfection, which were generally ineffective, destructive, or deleterious to filter performance (Price & Chu, 2020). These events highlight the need for institutions to address the gap in access to effective disinfection equipment for the reuse of N95 respirators which are needed to ensure the safety of the healthcare work force. (Bauchner, Fontanarosa, & Livingston, 2020; Livingston, Desai, & Berkwits, 2020) We show that ozone can be used to disinfect N95 respirators for multiple cycles without degrading filtration performance.

The Centers for Disease Control (CDC) recognized conditions under which the reuse of PPE, including N95 respirators, may be necessary to protect HCP and reduce the inherent risk of infection transmission at work. The CDC emphasized that their recommendations for extending the use of PPE were intended for healthcare institutions with professional respiratory protection programs. (U.S. Centers for Disease Control, 2020)

Four fundamental questions must be addressed when considering methods for reuse of PPE by healthcare workers:

1. Does the method effectively kill targeted organisms on the PPE?
2. Does the method degrade the function of the PPE?
3. Does the method create new dangers to healthcare workers?
4. Is the method practical in the current setting of the Covid-19 pandemic or similar future emergencies that may arise in areas with insufficient resources to maintain adequate supplies of PPE?

Several organizations published protocols based on previously published studies to address the shortage of N95 respirators (Applied Research Associates, 2020), including University of Nebraska School of Medicine (Lowe et al., 2020) and Duke University (Schwartz et al., 2020). Stanford University and 4C Air investigated N95 disinfection with dry heat at 75 degrees Celsius, showing that the filtration capability was not degraded below 95% for up to 20 cycles. (Cui et al., 2020) Another group based in Cleveland evaluated peracetic acid, demonstrating sterilization of spores on N95 masks. (John et al., 2020)

Workers at the National Institute of Allergy and Infectious Diseases evaluated the effectiveness of Vaporized Hydrogen Peroxide (VHP), UV-C light, dry heat, and ethanol. They showed that dry heat for 60 minutes can achieve a >99.9% reduction in SAR-CoV-2 (the virus responsible for COVID-19) and is suitable for up to two cycles of N95 disinfection on the basis of respirator fit testing. They also showed that UV-C and VHP can be used for up to 3 cycles of disinfection, and that ethanol is unsuitable because it damages filter performance. (Fischer et al., 2020)

Our group was interested in the feasibility of ozone disinfection of N95 respirators as an alternative for healthcare professionals and organizations that might not have access to a VHP or other disinfection devices. Ozone is an appealing disinfector because it is a strong oxidant. Viruses may be inactivated by ozone acting on the protein structure of a virus capsid or on viral nucleic acids. (Tseng & Li, 2006) Furthermore, ozone can be generated from air, can be quickly destroyed, and leaves no residue.

### N95 Filtering Facepiece Respirators

The N95 number refers to the U.S. National Institute for Occupational Safety and Health (NIOSH) classification of filtering respirators. The ratings describe the ability of the device to protect the wearer from dust and aerosolized liquid droplets in the air. The letter N indicates that a device is not resistant to oil and 95 refers to the minimum filtration efficiency percentage (95%) necessary for retaining 0.3 μm particulates.

3M Corporation (Minneapolis, MN) manufactures many N95 respirator models for industrial and healthcare applications. Table 1 summarizes the material composition of three models that are widely used in healthcare facilities. All have the same basic three-layered construction, wherein the middle layer has the smallest pores and an electrostatic field.

**Table 1.** 3M™ N95 Disposable Respirator Material Composition

| **Parts** | **1860, 1860S** | **1870+** | **8000 series** |
| --- | --- | --- | --- |
| Straps | Braided Polyisoprene | Polyisoprene | 8812, 8822 – Polyisoprene<br>8710, 8210 – Thermoplastic Elastomer |
| Staples | Steel | Steel | 8812, 8822 – Steel<br>8710 and 8210 – no staples |
| Nose Clip | Aluminium | Aluminium | 8210 – Aluminium<br>8710, 8812, 8822 – Steel |
| Nose Foam | Polyurethane Foam | Polyurethane | Polyester |
| Filter | Polypropylene/Polyester | Polypropylene/Polyester | Polyester/Polypropylene |
| Shell | Polypropylene | Polypropylene | na |
| Coverweb | Polypropylene | Polypropylene | na |
| Valve | na | na | Polypropylene |
| Valve diaphragm | na | na | Polyisoprene |

### Sterilization with Ozone

Ozone disinfectors are utilized in many applications, including water treatment. (Wojtowicz, 2000) Recently, continuous positive airway pressure (CPAP) mask disinfection systems utilizing gaseous ozone have been introduced to the market (without FDA approval), raising awareness of ozone disinfection but generating concern from the FDA due to reports of unexpected asthma attacks, headaches, and shortness of breath (U.S. Food and Drug Administration, 2020a).

Indeed, in the comments responding to the JAMA call for suggestions, contributors proposed using CPAP disinfection systems for N95 respirators (Bauchner et al., 2020) despite their lack of FDA approval. Some of these systems do not include an ozone monitor or destruction catalyst, thereby increasing the chance for hazardous exposure to users of the devices. Since ozone concentration levels above 5 ppm are considered immediately dangerous to life and health (U.S. National Institute of Occupational Safety and Health, 1994), ozone for disinfection or sterilization should be utilized only within systems designed to prevent hazardous exposure.

Foundational work by Ishizaki, et al. (Ishizaki, Shinriki, & Matsuyama, 1986) showed the impact of concentration, time, and humidity on the kill rate of bacterial spores. Below 50% relative humidity, gaseous ozone is not an effective disinfectant. However, the kill rate increases dramatically at a relative humidity of 80% or greater. For example, the kill rate of bacterial spores doubles when humidity is increased from 90% to 95%. Additional work by Sakurai, et al. examined the ozone kill rate as a function of the substrate material properties, including type (metal or type of polymer), pore size, chemical composition, surface roughness, and hydrophobicity. They also studied the kill rate for two different strains of the same organism. For a given ozone concentration and humidity level, the kill rate varied by up to a factor of 5. (Sakurai et al., 2003)

Two groups (Hudson, Sharma, & Vimalanathan, 2009) (Iwamura et al., 2013) demonstrated the feasibility of sanitizing hospital rooms using ozone, with Iwamura et al. finding that a concentration multiplied by exposure time (CT) of 25,000 ppm*min was required for room decontamination at the >log 3 level. Ozone has been shown to effectively kill sudden acute respiratory syndrome (SARS)-related coronavirus as early as 2004. (Zhang, Zheng, Xiao, Zhou, & Gao, 2004) Recently, researchers at Nara Medical University in Japan announced via press release that they had killed SARS-CoV-2 on a piece of stainless steel with a CT value of 330 (6 ppm * 55 min)(Nara Medical University, 2020) and humidity in the range of 60-80%.

The range of documented ozone concentrations needed to kill bacteria and viruses is 10,000 - 50,000 ppm*min, as shown in Table 2. High concentrations provide several benefits including mitigation of chemical depletion effects that could arise as ozone penetrates a pore, increased driving force for diffusion into the biofilm, and reduction of disinfection time. However, high concentrations present a more significant hazard to personnel operating the equipment and a higher rate of reaction with materials of construction. Therefore, the choice of concentration and time should be optimized for the application.

**Table 2.** Summary of Conditions Used for Testing Disinfection of Materials. **Legend:** O_3_ = ozone; O_2_ = oxygen; CT = concentration multiplied by exposure time; ppm = parts per million; RH = relative humidity; Time = time of exposure; Kill (log) = “kill efficiency” expressed as the base 10 logarithm of the reduction in organisms. Log 3 = 99.9%, log 6 = 99.9999%, etc.; atm = atmosphere. ^^^Assumes room temperature and pressure (RTP) (25 deg. C.,1 atm, ideal gas approximation)

| Group | Target | O<sub>3</sub><br>mg/L | O<sub>3</sub><br>ppm<sup>^</sup> | RH | Time | Kill<br>(log) | CT<br>ppm*min | Comment |
| --- | --- | --- | --- | --- | --- | --- | --- | --- |
| Bedard et al., 2009 | not specified | 85 | 43208 | 95% | 120 | 12 | 5185000 | ~0.5 atm, ~10% O3/O2 mix |
| de Souza Botelho-Almeida et al., 2018 | *Geobacillus stearothermophilus* ATCC 7953 | 7 | 3558 | >80% | 80 | 6 | 284667 | O3/O2 mix ~1.2 atm |
| Hudson et al., 2009 | H3N2 influenza virus | 0.04 | 20 | 38% | 20 | 0.1 | 400 | Ozone / Air room sterilization (various room sizes / surfaces) |
|  |  | 0.04 | 20 | 70% | 20 | 2.6 | 400 |  |
|  |  | 0.06 | 28 | 95% | 60 | 2.1 | 1680 |  |
| Ishizaki et al., 1986 | *Bacillus subtilis* NCTC 10073 | 1.00 | 508 | 90% | 360 | 6 | 183000 | Ozone / Air |
| Iwamura et al., 2013 | *B. atrophaeus* ATCC 9372 | 0.43 | 200 | >80% | 120 | 3 | 24000 | Ozone / Air |
| Nakamura, 2008 | *Pseudomonas aeruginosa* | 0.10 | 51 | agar | 120 | 6 | 6100 | Agar plate |
|  | H1N1 influenza virus | 0.02 | 10 | 65% | 210 | >4 | 2100 | Ozone/Air in 160L chamber |
|  |  | 0.04 | 20 | 65% | 150 | 5 | 3000 |  |
|  |  | 0.04 | 20 | 65% | 600 | 6 | 12000 |  |
|  |  | 1.29 | 43208 | <50% | 40 | none | 24000 |  |
|  |  | 0.86 | 3558 | ~80% | 120 | >3 | 48000 |  |
| Nara Medical University | SARS-CoV-2 |  | 6 | 60-80% | 55 | >3 | 330 | Stainless steel substrate |

### Potential Ozone Damage to N95 Respirators

The authors are not aware of previous studies on potential ozonation damage to N95 respirators or polypropylene non-woven electret filters under conditions typically employed in disinfection with ozone gas. There is a recent study of the impact of ozone on polypropylene electret non-woven filter material. This study found very little impact on the filtration efficiency of the filter material subjected to ozone doses of 200 ppm for 90 minutes and 20 ppm for 36 hours at room temperature and a relative humidity level of 55%.(Dennis, Pourdeyhimi, Cashion, Emanuel, & Hubbard, 2020) Although this result is encouraging, it is not definitive, since it was not conducted at a humidity level or ozone concentration typically associated with >log 2 disinfection by ozone gas. (Ishizaki et al., 1986)

In theory, polypropylene (the filter material) should have good chemical compatibility with ozone because it lacks double bonds or functional groups that react strongly with ozone. (Rogers, 2012) However, as shown above in Table 2, ozone gas requires high humidity to be effective for sterilization (typically ≥80%), and humidity has been shown to reduce the electrostatic potential of electret materials. (Motyl & Lowkis, 2006)

Fortunately, a recent study showed that the practical impact of humidity on polypropylene electret filters can be negligible. (Lee & Kim, 2020) The investigators showed that the filtration performance of polypropylene electret filters did not degrade after 48h at 90% relative humidity at room temperature, even though the electrostatic potential was reduced 11-12%. They found that the impact of humidity on electret materials is correlated with the wettability of a material as measured by the contact angle of water droplets on the material. Further confirmation that humidity levels at or below 90% do not seem to impact N95 filters is provided by a study of N95 masks aged past their stated shelf life. N95 filters that had been stored for six years in a warehouse with humidity levels varying from 20 to 85% were shown to perform within specifications. (Viscusi, Bergman, Sinkule, & Shaffer, 2009)

In contrast to the filter media, the elastic band or other components of respirators could be susceptible to ozone attack if made from natural or synthetic rubber, which contains unsaturated carbon-carbon bonds. (Rogers, 2012) Ultimately, the relative rates of viral destruction versus polymer / electret degradation by ozone will determine whether ozone disinfection is effective. The best success criteria are functional testing of filtration efficiency and functional human fit testing. (Fischer et al., 2020)

Therefore, our aims are:

1. to assess three factors that are critical for ozone to effectively kill targeted organisms on N95 respirators for disinfection and reuse: ozone concentration, humidity, and length of disinfection treatment;
2. to determine the effect of ozone at similar (to number 1) concentrations, humidity, and times on the integrity and filtration rate of N95 respirators;
3. to review the safety of ozone-based disinfection technology in relation to potential risks or dangers to HCP or to the environment and to compare ozone safety to other methods currently in use;
4. to assess the feasibility and practicality of this ozone-based disinfection.

## METHODS

### Respirators

Three different N95 respirators were tested: 3M 1860, 3M 1870, and 3M 8000. These respirators were chosen because they are being widely used in hospitals during this pandemic.

### Prototype: Ozone Disinfection Test Equipment

Based on review of the literature, the authors estimated that concentration-times of 50,000 ppm*min or less in humidity ≥ 80% could potentially kill bacteria on N95 respirators (see Table 2 above). A prototype ozone chamber produced by Ozone Solutions (Iowa, USA) was used for testing the efficacy of ozone for killing bacteria on N95 respirators. The ozone chamber (see Figure 1) components included: airtight chamber, ozone generator, ozone destruct unit, and ozone UV analyzer. The chamber can concentrate ozone up to 500 parts per million (ppm) and precisely record ozone concentration using an ultraviolet ozone analyzer. The ozone generator (HP-200) produces ozone using a small pump to draw in ambient air at a rate of 1-2 liters per minute. The ozone generator produces ozone and maintains the ozone level in the sealed chamber at a fixed concentration based on the analyzer measurements. The chamber’s ozone destruction catalyst breaks down ozone in the chamber at a rate of 40 ppm/min, allowing the user to open the chamber safely without threat from elevated ozone levels after a modest time interval.

**Figure 1.**
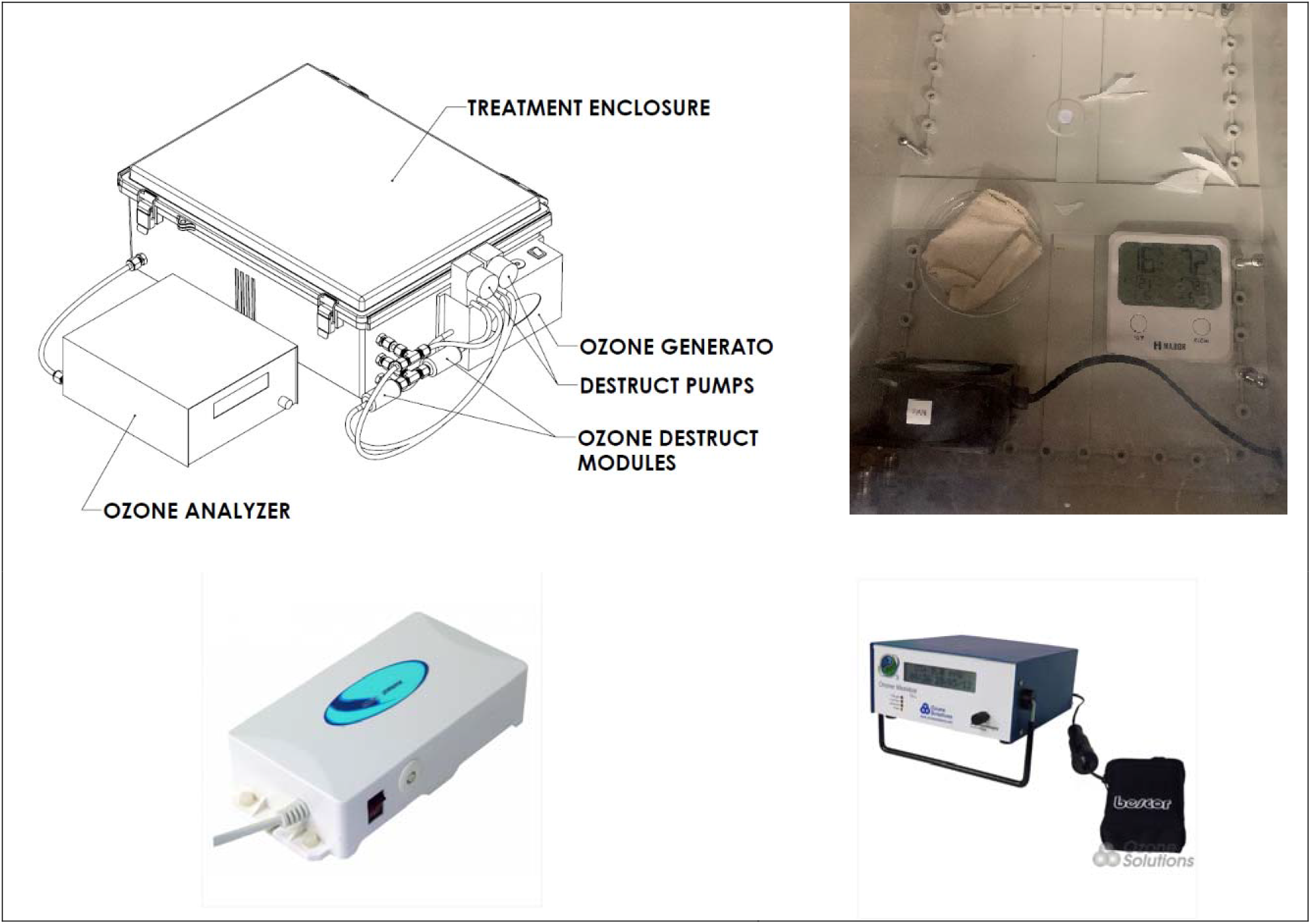
Prototype: Ozone Disinfection Test Equipment. **Legend:** Ozone Solution’s prototype of self-contained ozone chamber. Sealable chamber dimensions: 9” high × 30” deep × 20” wide. Top left: prototype ozone chamber inside fume hood for kill efficacy testing. External ozone analyzer connects to sealed chamber. Top right: inside of sealed chamber, fan at bottom left for ozone circulation, glassware and paper for wicking water to maintain high ambient humidity, hygrometer/thermometer to measure ambient conditions within the chamber during testing. Bottom left: HP-200 (110V/60Hz) low cost ozone generator with air pump. Right: UV-106L ozone concentration analyzer.

### Ozone Kill Efficiency Testing

Four pieces of approximately 4 square centimeters (sq cm) from each type of N95 respirator (3M 1860, 3M 1870, and 3M 8000) were dipped in *Pseudomonas aeruginosa* (PsA) containing bacterial broth (PA01 culture). PsA was chosen because it is a vegetative bacteria with high innate resistance to chemical germicides. According to CDC, vegetative bacteria such as PsA are more resistant than lipid viruses to chemical germicides and sterilization processes.(Rutala, 2008) Therefore, SARS-CoV-2 virus (responsible for COVID-19) is likely to be more susceptible to conditions that prove effective at killing PsA. Samples of the 1860 masks including the metal nose clip were also included.

Half of the respirator pieces were placed inside the ozone chamber and exposed to 400 ppm at 80% relative humidity for two hours at room temperature. The remaining half of the pieces were kept in ambient air with approximately 35% humidity for two hours. The N95 pieces were then dipped in 1 milliliter (ml) Phosphate Buffer Solution (PBS) and vortexed hard. Serial dilutions of this solution were plated and incubated overnight in a 37°C incubator. Number of bacteria were enumerated by counting the colonies in dilutions up to 1×10^7^. Bacterial growth was calculated as colony-forming units per milliliter (CFU/ml). Kill efficacy was calculated by comparing the number of CFU/ml of the ozone-exposed respirator culture compared to the ambient air-exposed controls.

### Ozone Effect on Filtration Efficiency Testing

The 1860 and 1870 respirators were tested for the effects of ozone on filtration efficiency at conditions similar to those used for testing ozone’s kill efficacy. Respirators were exposed to 450 ppm ozone for two hours at 75-90% humidity for multiple cycles to determine the effects of repeated exposures as shown in Table 3. Between each cycle, respirators were exposed to room air at ambient conditions for at least twenty minutes. All ozone exposures were performed at Ozone Solutions (Iowa, USA).

The effects of ozone treatment were tested on four 1860 respirators by 4C Air (California, USA) (See Table 3.). All samples were tested for initial filtration properties via a standard filtration efficiency test used in NIOSH N95, 42 CFR Part 84 (Respiratory Protective Devices). Tests were conducted on an “Automated Filter Tester” 8130A (TSI, Inc.) using 0.26 μm (mass median diameter) aerosolized sodium chloride under a flow rate of 85 L/min, the industry standard equipment for aerosol filtration efficiency testing. (Lore, Sambol, Japuntich, Franklin, & Hinrichs, 2011)

The effects of ozone treatment were tested on 15 1870 respirators by the CDC as shown in Supplemental Section D. (U.S. National Personal Protective Technology Laboratory, 2020) Five samples were tested using a modified version of the NIOSH Standard Test Procedure (STP) TEB-APR-STP-0059 to determine particulate filtration efficiency. The TSI, Inc. model 8130 using sodium chloride aerosol under a flow rate of 85 L/min was used for the filtration evaluation. The Instron® 5943 Tensile Tester was used for this evaluation. Five samples were tested using a Manikin Fit Factor with Statis Advanced Headform (Hanson Robotics). Fit factor is a quantitative fit testing protocol as defined by OSHA 1910.134(f)(7) wherein tight-fitting facepieces undergo a pass/fail test to assess change in fit performance associated with disinfection of respirators. Scores range from zero to 200. Scores greater than 100 are considered to have passed this quantitative fit testing on a mannikin headform, i.e., no change in fit performance is detected. Tests were performed on stationary mannikin headforms simulating normal and deep breathing. Five samples were used as controls for these tests. Additionally, tensile strength testing of the straps was performed to determine percent change in strap integrity as defined by the tensile strength of exposed respirators minus the controls divided by the controls. The full CDC assessment plan can be found at: https://www.cdc.gov/niosh/npptl/respirators/testing/pdfs/NIOSHApproved_Decon_TestPlan10.pdf.

**Table 3.** Number of Exposure Cycles for Respirators Tested for Effects of Ozone on Filtration Efficiency at 450 ppm ozone with 75-90% humidity for 2 hours per cycle. **Legend:** Data on the effects of different numbers of exposure cycles were collected at two sites: 4C Air and CDC. Both locations used the same test conditions: 450 ppm ozone at 75-90% humidity for 2 hours per cycle.

|  |  |  |
| --- | --- | --- |
| **Mask type** | 1860 respirators | 1870 respirators |
| **Control** | Data from 4C Air | Data from CDC |
| **3 cycles** | 2 respirators |  |
| **5 cycles** |  | 10 respirators |
| **10 cycles** | 2 respirators |  |
| **Sample size** | **4 respirators** | **10 respirators** |

### Statistical analysis

Comparisons between ozone-exposed and control groups were made using a two-tailed Student’s T-test with threshold for significance defined as *P* < 0.05.

## RESULTS

Exposure to ozone at 400 ppm with 80% humidity for two hours effectively killed bacteria on N95 respirators, types 1860, 1870, and 8000, as shown in Figure 2 and Table 3.

### Ozone Kill Efficacy

The data on bacterial growth from respirator pieces are summarized in Table 3 with log kill efficacy. Control respirator pieces that were exposed to air, but not to ozone, grew log 8.4 to 9.6 colony-forming units (CFU) per respirator piece. Respirators exposed to ozone 400 ppm 80% humidity for two hours (ozone-treated) yielded little to no growth (zero CFU/ml), with the exception of one sample from the 1860 respirator (sample 2.1). Pictures of bacterial growth from the ozone-treated respirator pieces and controls are shown in Figure 2.

**Figure 2.**
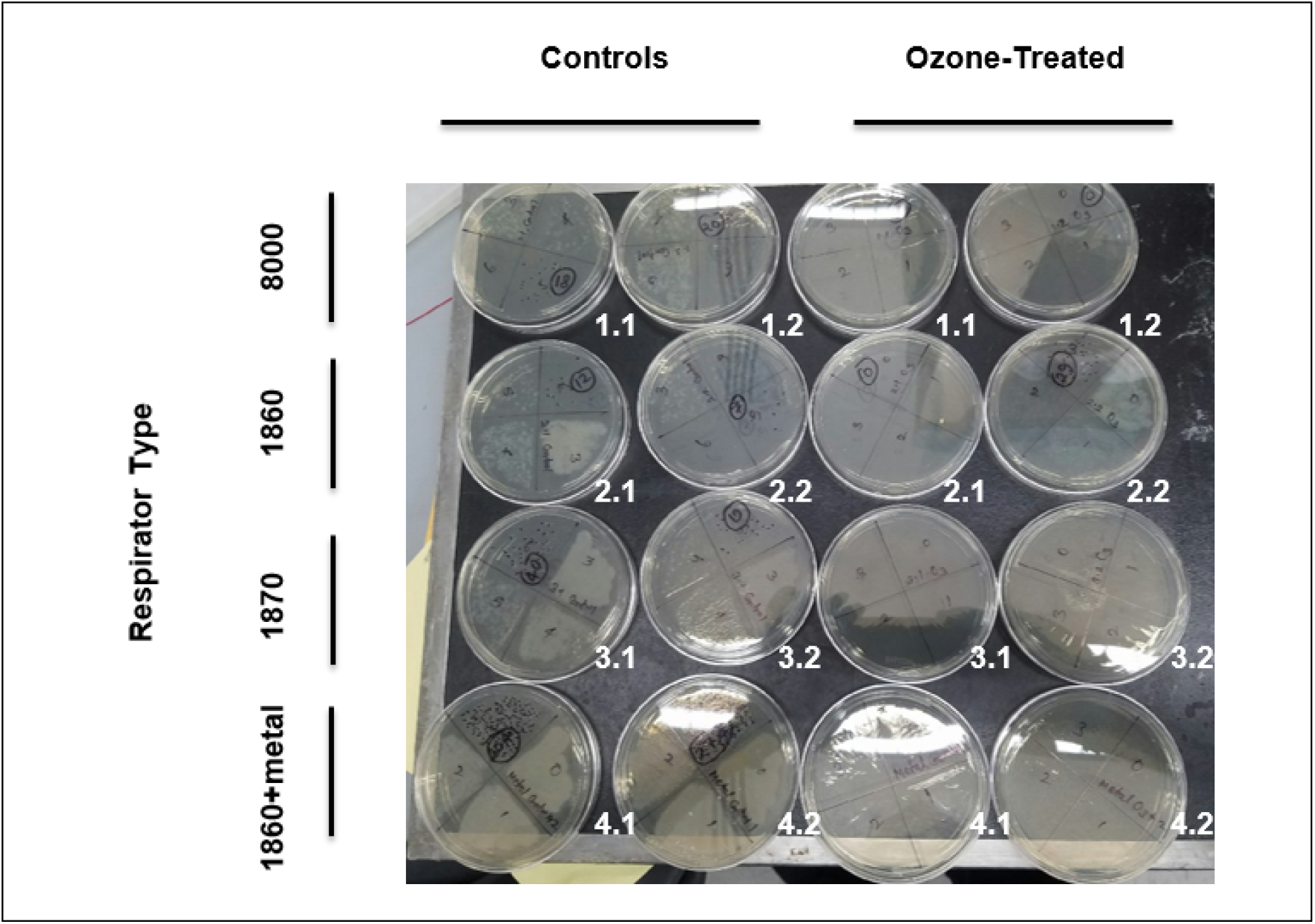
Ozone-treated and Control Respirator Sample Cultures Inoculated with Bacteria. **Legend:** Control columns: cultures from respirators inoculated with bacterial culture, exposed to ambient air 35% humidity for two hours, and incubated for 24 hours. Ozone-treated columns: cultures from respirators inoculated with bacteria culture, exposed to 400 ppm ozone 80% humidity for two hours, and incubated for 24 hours. Rows are labeled to identify respirator types tested. Tests were performed in duplicate for each respirator type. Serial dilutions were performed to enumerate the numbers of live bacteria.

**Table 4.** Log Kill Efficacy in Ozone-Exposed Respirator Pieces compared to Controls. **Legend**.CFU = colony-forming units; ND = Not detected. Kill yield log is expressed as the base 10 logarithm of the reduction in organisms. Log 3 = 99.9%, log 6 = 99.9999%, etc.

| Respirator | Case number | Log CFUs/respirator piece (Control group) | Log CFUs/respirator piece (Ozone group) | Kill yield Log (%) |
| --- | --- | --- | --- | --- |
| 1870 | 1.1 | 7.95 | ND | >7 |
|  | 1.2 | 8.00 | ND | >8 |
| 1860 | 2.1 | 8.78 | ND | >8 |
|  | 2.2 | 7.54 | 6.16 | 1.38 |
| 1860 + Nose Clip | 4.1 | 7.01 | ND | >7 |
|  | 4.2 | 7.14 | ND | >7 |
| 8000 | 3.1 | 9.30 | ND | >9 |
|  | 3.2 | 8.98 | ND | >9 |

### Ozone Effects on N95 Function

Respirator filtration efficiency tests revealed that all respirators maintained > 95% efficiency (the filtration standard to maintain N95 function) after exposure to ozone under various conditions and multiple cycles, as shown in Table 4. There was no significant change in filtration efficiency (*P* = 0.45) between ozone-exposed 1870 respirators and their controls. There was no significant change in filter resistance (*P* = 0.84) between ozone-exposed 1870 respirators and controls. There was little to no noticeable wear to the 1860 or 1870 respirators after 10 cycles of 450 ppm ozone for two hours at 75-90% humidity (see Supplemental Section D). Ozone treatment was 450 ppm for 2 hours at 75-90% humidity.

Quantitative fit testing using mannikin headforms revealed no loss of fit after ozone exposures. All respirators exposed to ozone passed quantitative fit testing as defined by OSHA 1910.134(f)(7) with little difference compared to control respirators. Ozone had variable effects on the respirator straps. The integrity of the N95 1870 respirator straps did not appear to suffer from similar ozone exposure. No visible degradation of the straps was observed, and there was minimal reduction in tensile strength of 1870 straps, 0.2% (*P* = 0.99) for the top strap and 5.3% (*P* = 0.65) for the bottom strap as shown in Table 5. In contrast, elastic bands on the 1860 respirators were negatively affected during ozone exposure with visible damage and cracking. There was a residual odor on the respirators after ozone exposure.

**Table 5.** N95 1860 Filtration Testing after Exposure to Ozone 450 ppm at 75-90% humidity for 2 hours per cycle. **Legend**. 1860 N95 respirators samples were exposed to ozone at Ozone Solutions (Iowa, USA) and filtration tests were performed at 4C Air (California, USA). 1870 N95 respirators were exposed to ozone at Ozone Solutions (Iowa, USA) and filtration tests were performed at the CDC (Pittsburgh, PA). 5A (Top): Filtration efficiency testing. All masks were tested to determine the effects of ozone on filtration efficiency using the TSI, Inc. model 8130 (machine tested). There were inadequate sample sizes to calculate SEM for measurements on the 1860 respirators. 5B (Middle): Mannikin Headform Quantitative Fit Testing. 5C (Bottom): Strap Integrity Testing. There was no significant reduction in tensile strength of 1870 straps as a result of ozone exposure, 0.2% for the top strap (*P* = 0.99) and 5.3% for the bottom (*P* = 0.65).

| **Respirator Type** | **Number of Treatment Cycles** | **Mean Filtration Efficiency ± SEM (%) (range)** | **Mean Filter Resistance ± SEM (mmH<sub>2</sub>O) (range)** |
| --- | --- | --- | --- |
| **1860 N95 Respirator (Machine tested) n=2** | **0 (Controls)** | 99.79 | 8.8 |
|  | **3** | 99.70 | 8.0 |
|  | **10** | 99.64 | 7.9 |
| **1870 N95 Respirator (Machine tested) n=5** | **0 (Controls)** | 99.42 ± 0.43<br>(99.57-99.87) | 8.53 ± 0.18<br>(8.2-8.6) |
| | **5** | 99.83 ± 0.04<br>(99.72-99.93) | 8.64 ± 0.45<br>(7.8-10.3) |

**5B: Mannikin Headform Quantitative Fit Testing**
| **Respirator Type** | **Number of Treatment Cycles** | **Mean Normal Breathing 1 ± SEM (mmFF) (range)** | **Mean Deep Breathing ± SEM (mmFF) (range)** | **Mean Normal Breathing 2 ± SEM (mmFF) (range)** | **Mean Overall Fit Factor ± SEM (mmFF) (range)** |
| --- | --- | --- | --- | --- | --- |
| **1870 N95 Respirator (Machine tested) n=5** | **0 (Controls)** | 194 | 161 | 162 | 169 |
| | **5** | 190 ± 9 | 190 ± 18 | 180 ± 14 | 180 ± 14 |

| **Respirator Type** | **Number of Treatment Cycles** | **Top Strap Mean Force ± SEM (N) (range)** | **Bottom Strap Mean Force ± SEM (N) (range)** |
| --- | --- | --- | --- |
| **1870 N95 Respirator (Machine tested) n=3** | **0 (Controls)** | 1.708 ± 0.083<br>(1.581-1.865) | 1.753 ± 0.028<br>(1.703-1.799) |
| | **5** | 1.704 ± 0.222<br>(1.263-1.978) | 1.660 ± 0.175<br>(1.313-1.876) |

## DISCUSSION

This study demonstrates that an ozone application achieves a high level of disinfection against PsA, a vegetative bacteria that the CDC identifies as more difficult to kill than medium sized viruses such as SARS-CoV-2 (Covid-19). (Rutala, 2008) Furthermore, conditions shown to kill these bacteria did not damage or degrade respirator filtration. This is the first report of successful disinfection of N95 PPE with ozone of which the authors are aware. It is also the first report, to the authors’ knowledge, to identify necessary conditions for ozone to kill organisms on N95 masks without degrading the function of N95 filters.

### Effectiveness of Ozone for Disinfection of N95 Respirators

Ozone disinfection is an effective method for killing bacteria from contaminated N95 respirators commonly used in healthcare settings during the current COVID-19 pandemic. The conditions for 99+% kill efficacy in this study were: ozone concentration at or above 400 ppm with 80% humidity for two hours. Experiments in our lab with similar concentrations of ozone and humidity below 50% did not yield kill (refer to supplementary data). Other literature supports the need for humidity.(Ishizaki et al., 1986) The necessary conditions that we have identified to kill bacteria on N95 respirators fall within the range of conditions used for other applications of ozone sterilization in the literature as shown in Table 2. Lower concentrations of ozone for shorter periods of time may achieve similar levels of efficacy on viruses based on previous work with influenza virus.(Tanaka, Sakurai, Ishii, & Matsuzawa, 2009) Further testing is necessary to determine if the conditions used in this experiment can be optimized.

For 1870 and 8000 N95 respirators, we achieved >log 6 kill in two sets of experiments. For 1860 masks, in each experiment, we achieve > log 6 kill for at least one sample, but failed to disinfect at least one sample. Additional work is required to understand the cause of the failure but this might be an artifact of our experimental method, which utilized samples cut from a mask. In the case of the 1860 masks, the layers separated from each other, and it seems possible that bacteria penetrated the interlayer space where they were protected somewhat from the ozone.

### Effects of Ozone on Filtration and Integrity of N95 Respirators

Ozone did not degrade the function or fit of the respirator filters after ten cycles of ozone at 450 ppm at 75-90% humidity for two hours at room temperature. The filtration portion of N95 respirators consist primarily of polyethylene and polypropylene. The elastic bands are latex rubber which is known to be easily damaged by ozone, and the nose foam is made from polyurethane, which does not exhibit good ozone compatibility. (Rogers, 2012) A lack or decrease in nose foam does not affect filtration efficiency of N95 respirators, though it may affect fit and comfort to the wearer. However, no ill effects on fit were measured during the quantitative fit testing of 1870 respirators. There was also no significant decrease in the tensile strength of the 1870 straps when compared to controls. This suggests that ozone does not affect the fit of the 1870 masks. The source of the residual odor is unclear and may result from reactions of ozone with materials in the chamber other than the respirator filtration components, such as the bacteria itself, glassware, or the moist paper. This is being investigated.

### Comparison of Ozone to Other Disinfection Systems Given FDA Emergency Use Authorizations

In the past seven weeks, the FDA issued five emergency use authorizations (EUAs) to allow decontamination of N95 compatible respirators for re-use by healthcare personnel. Each of these disinfection systems uses vaporized hydrogen peroxide (VHP). The five systems are:

1. The Battelle Decontamination System (EUA issued 3/29/20) uses VHP for up to 20 cycles per respirator. (U.S. Food and Drug Administration, 2020d) Each large, self-contained chamber has a maximum capacity to disinfect 10,000 respirators per load. This technology is based on previous work conducted under contract to the FDA demonstrating log-6 kill.(Battelle Memorial Institute, 2016)
2. The STERIS Sterilization Systems (EUA issued 4/9/20) uses VHP for up to 10 cycles per respirator.(U.S. Food and Drug Administration, 2020f) Each unit has a maximum capacity to disinfect 10 respirators per load.
3. The ASP STERRAD Sterilization System (EUA issued 4/11/20) uses VHP for up to 2 cycles per respirator. (U.S. Food and Drug Administration, 2020c) Each unit has a maximum capacity to disinfect 10 respirators per load.
4. The Stryker STERIZONE VP4 N95 Respirator, which utilizes ozone is used as an adjunct to VHP for Decontamination Cycle (EUA issued 4/14/20), for up to 2 cycles per respirator. (U.S. Food and Drug Administration, 2020g) Each unit has a maximum capacity to disinfect 20 respirators per load
5. Sterilucent HC 80TT Hydrogen Peroxide Sterilizer (EUA 4/20/20) for up to 10 cycles per respirator. (U.S. Food and Drug Administration, 2020e) Each unit has a maximum capacity to disinfect 12 respirators per load.
6. Duke Decontamination System (EUA 5/7/20) for up to 10 cycles per respirator. (U.S. Food and Drug Administration, 2020b) The system uses VHP. This system is operated in five rooms at Duke University affiliated hospitals, with capacities of 882 to 1764 respirators per batch.

Ozone appears to be as effective at killing bacteria on N95 respirators as the five FDA approved methods. Ozone does not impair the filtration function of the N95 respirators after 10 or more cycles, exceeding maximum cycles for two of the four VHP systems that have received FDA EUAs. Preliminary fit testing showed no significant change following ozone disinfection. Based on the results to date, ozone disinfection appears to be a viable alternative to current VHP approaches that have received FDA EUA.

### Safety to Healthcare Personnel and the Environment

This application of ozone appears safe. The ozone chamber in this experiment contained an analytical device to accurately measure ozone levels and an ozone destruction catalyst that enables rapid destruction of ozone in a matter of minutes. This combination provides reasonable safety for ozone disinfection. Ozone chambers can be easily manufactured to safely and accurately reproduce and monitor the internal environment for disinfection of N95 respirators. It is critical that ozone application equipment contain these monitoring and ozone destruction capabilities to ensure safety during disinfection.

After a short post-processing period, ozone reverts back to oxygen thereby posing no concerns related to safety, in contrast with other methods. While ETO is a well-established, FDA-approved, commercialized sterilization technique, this method of disinfection of N95 masks cannot be practiced in clinical settings because of the hazards of ETO. Further, because of environmental concerns, ETO sterilization requires emission control equipment. VHP processes are effective, but there is concern that residual H2O2 will off-gas while PPE is worn, and therefore respirators disinfected by H2O2 must be checked to insure that off-gassing is complete. (Schwartz et al., 2020) Ozone lacks the environmental emissions or occupational exposure to the wearer associated with these methods of PPE disinfection.

In comparison to VHP and ETO disinfection methods, ozone appears to be a safer alternative for disinfection of PPE. While ozone is toxic at levels above OSHA standards, (U.S. National Institute of Occupational Safety and Health, 1994) the half-life of ozone to oxygen degradation can be very short under proper conditions, such as contact with water or a catalyst. (Wojtowicz, 2000) Ozone in the concentrations used here to kill PsA was catalytically converted back to oxygen in minutes. Ozone monitoring and destruction devices are necessary to ensure safety of personnel operating the device.

### Feasibility and Practicality of Ozone Disinfection

Some devices using ozone for sterilization require dedicated supplies of pure oxygen; however, ozone generators without dedicated oxygen supplies have the capacity to produce concentrations equal to those used during our tests. Therefore, it is reasonable to assume that the application of ozone is feasible in smaller medical facilities or offices without dedicated oxygen supplies or systems. Small ozone disinfection devices may be a cost-effective, easy-to-distribute solution for rural hospitals, clinics, and professional offices when N95 respirators must be reused due to insufficient supplies or lack of access to large decontamination facilities.

Limitations of these studies include the small number of samples tested, the limited number of ozone concentrations tested, and the use of only one infectious agent, PsA. Further investigations will test the lower limits of conditions, including ozone concentration (ppm), humidity, and time needed to effectively kill microorganisms. Larger samples and additional infectious agents should be tested. Additional strap testing and face fit are also needed. Further testing and confirmation of our results is necessary before widespread implementation.

## CONCLUSION

We have shown that ozone is an effective stand-alone method for decontaminating N95 respirators by killing bacteria at a level that is proven to also kill viruses. In addition, exposure to ozone concentrations of 400 ppm with relative humidity of 75-90% at room temperature, for up to 10 cycles of 2-hour treatments did not degrade N95 filtration. Moreover, ozone presents minimal risk to healthcare personnel or to the environment when used in professionally constructed sealed chamber devices with ozone monitoring and destruction equipment. Ozone disinfecting devices may provide a practical means of decontaminating N95 respirators especially for rural areas, healthcare personnel, and institutions that do not have access to large-scale disinfection facilities. Small ozone decontamination devices are practical and ozone generators are already available for other purposes. Existing guidelines for institutional protocols to decontaminate N95 respirators can be tailored for ozone devices. Ozone disinfection could augment other methods for PPE reuse, such as in combination with UV-C disinfection, with VHP, or with ETO to reduce off-gas time or remove toxic residuals. Before widespread adoption, further studies are needed to determine if the conditions used in these experiments can be optimized. Ozone decontamination may be an effective tool to extend the life and usage cycles for N95 respirators during the current pandemic and in future crises.

## Data Availability

All data referred to in this manuscripts has been disclosed in the main or supplementary text. Addition data found to be missing may be requested from the corresponding author.

## Acknowledgements

Erica L Herzog, Yale University

Jimmy Moler, Ozone Solutions

Kevin York, Ozone Solutions

Munroe Chirnomas, Corvalsys, Morristown, NJ

Alexandra Stephens, Smith College, Northampton, MA

Yi Cui, Stanford University, Palo Alto, CA

Lei Liao, 4C Air, San Jose, CA

Foad Katirai

Shigeru Saito This work is dedicated to health care workers and first responders who are fighting the COVID-19 pandemic.

EPM was funded by NHLBI Training Grant T32HL007778

LS was funded by ATS Critical Care Unrestricted Grant 2018

Prototype ozone disinfection chamber was provided by Ozone Solutions (Hull, IA)

## Supplemental Infromation

### Supplemental Section A

FDA Emergency Use Authorizations (EUA) as of 4/30/2020:

1. Battelle EUA available at: https://www.fda.gov/media/136529/download
2. Steris EUA available at: https://www.fda.gov/media/136843/download
3. ASP STERRAD EUA available at: https://www.fda.gov/media/136884/download
4. See FDA: Emergency Use Authorization (EUA) - EUA information and list of all EUAs - STERIZONE VP4 EUA available at: https://www.fda.gov/media/136976/download
5. Sterilucent HC 80TT Hydrogen Peroxide Sterilizer https://www.fda.gov/media/137169/download
6. Duke Decontamination System https://www.fda.gov/media/137762/download

### Supplemental Section B

Qualitative Trial Results

- Bacterial suspension of Pseudomonas aeruginosa (PsA)
- Small portion of respirator used (approximately 4 sq cm)

- Respirator portions 1-3: contaminated with PsA, exposed to ozone

- respirators portions 1, 2 and 3 were exposed to ozone
- Respirator portion 4: negative control (no contamination), exposed to ozone
- “Dry control” = portion of respirator contaminated with PsA, not exposed to ozone
- Concentration of >400 PPM for at least 2 hours (single cycle) with humidity greater than or equal to 80%
- “New control” = positive control, inoculation on petri dish with suspension of PsA, no ozone treatment

### Supplemental Section C

Initial attempts at bacteria kill

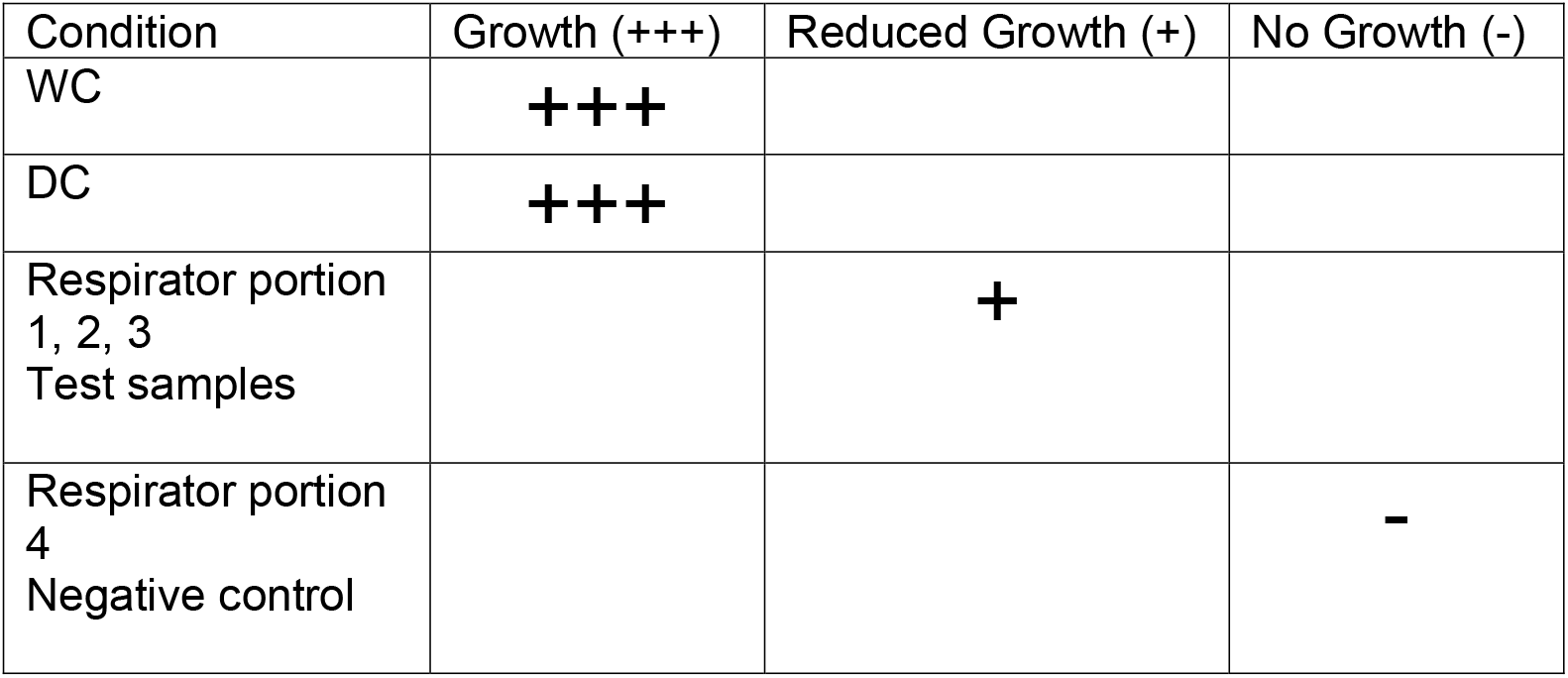

Attempt 3 images:

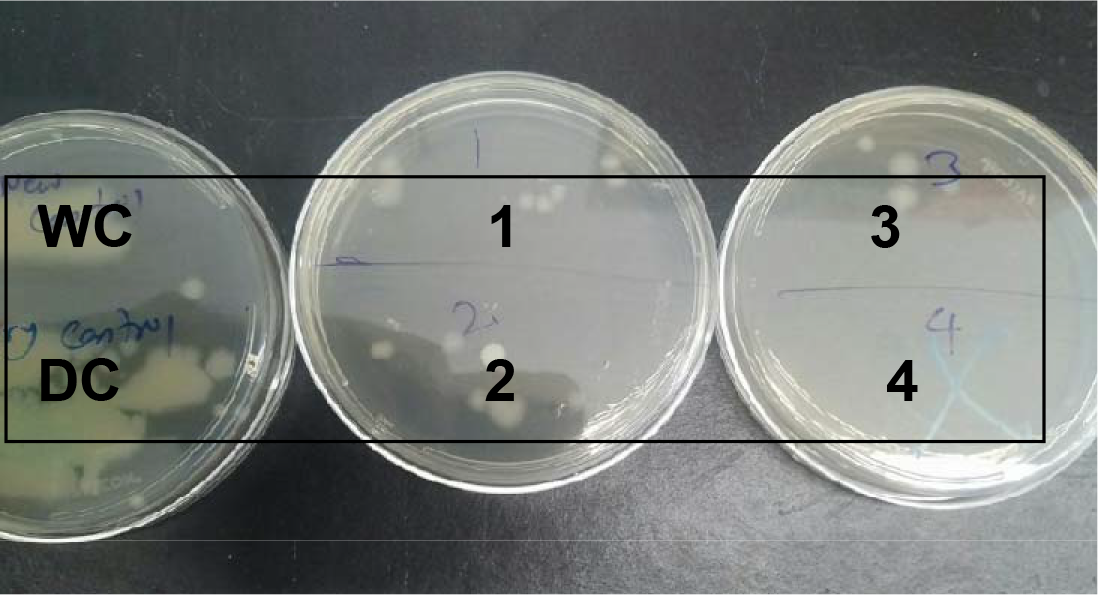

WC = PsA directly to dish

DC = PsA on respirator, left in ambient air, swab from respirator applied to dish)

1,2,3 = PsA on respirator, exposed to humid ozone chamber, swab from respirator applied to dish

4 = portion of respirator exposed to humid ozone chamber, swab from respirator applied to dish

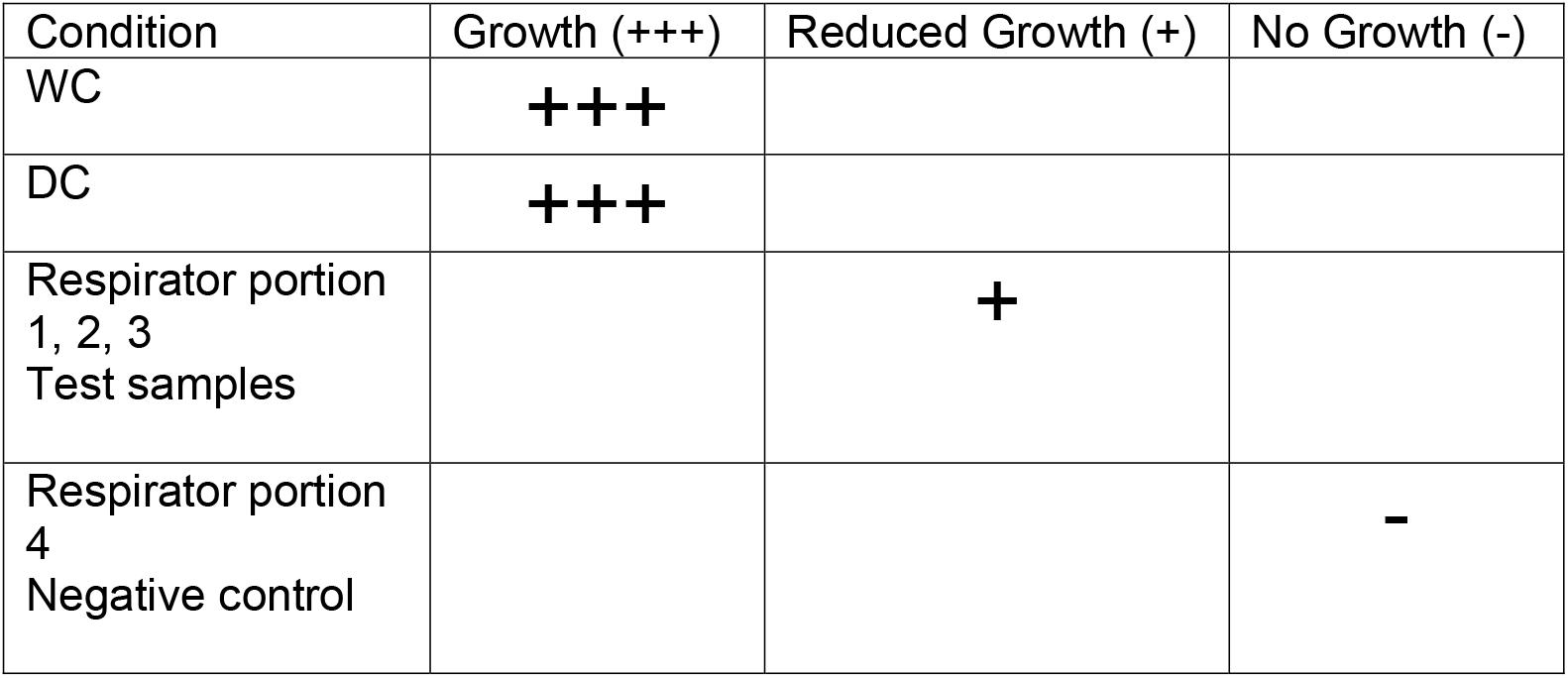

### Attempt 3 Ozone Kill Efficacy

Control respirator pieces that were exposed to air, but not to ozone, grew log 8.4 to 9.6 colony-forming units (CFU) per respirator piece as shown in Table 3. Respirators exposed to ozone 400 ppm 80% humidity for two hours (ozone-treated) yielded little to no growth (zero CFU/ml) as shown in Table 3, with the exception of one sample from the 1860 respirator (sample 2.1). Representative pictures of bacterial growth from the ozone-treated respirator pieces and controls are shown in Figure 2. Samples in Figure 2 were diluted 1, 10, 100 or 1,000 times denoted as 0,1, 2, and 3 respectively on the agar plates covers. The data on bacterial growth from respirator pieces are summarized in Table 3 with log kill efficacy.

**Figure C1.**
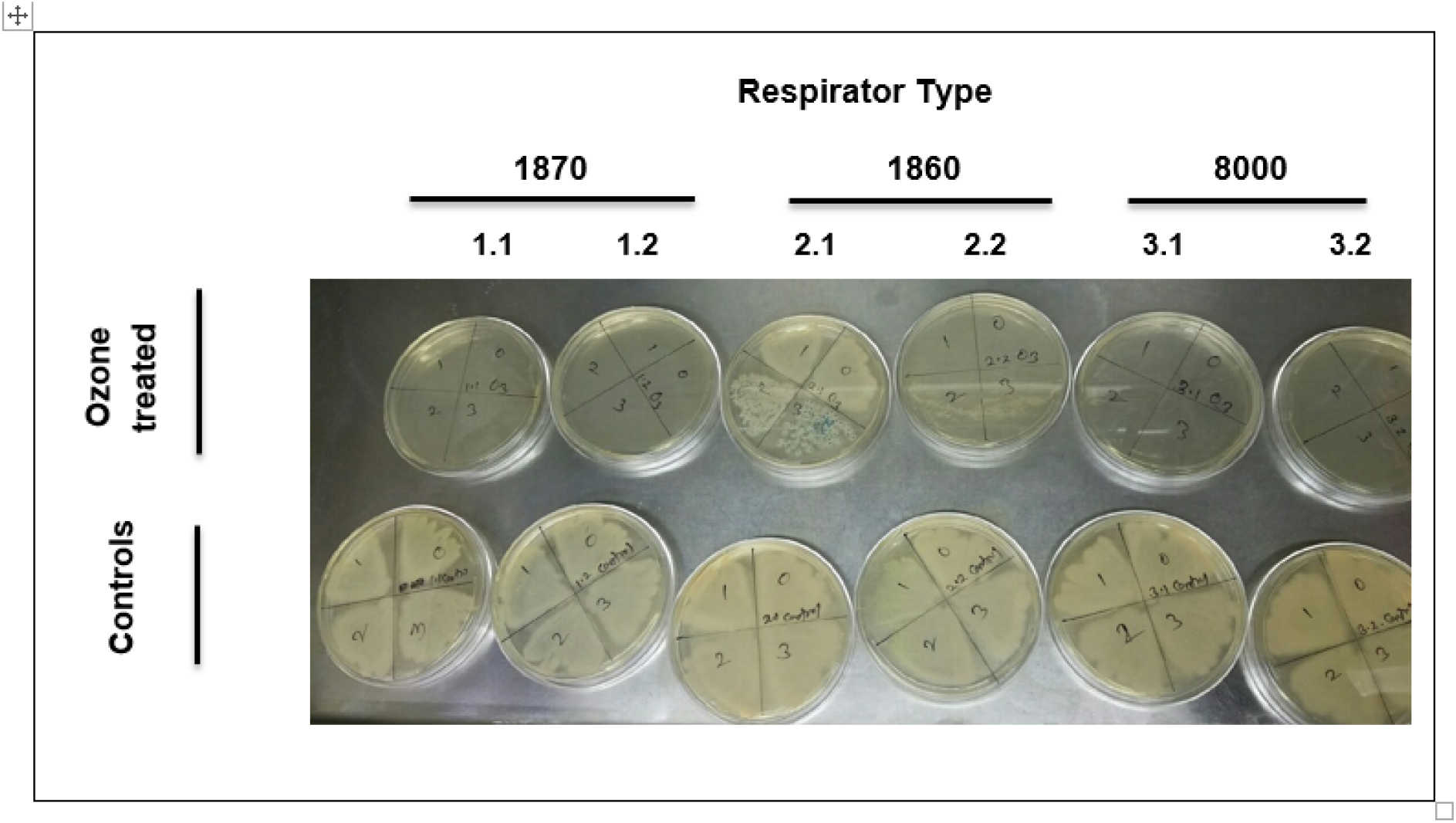
Cultures from Ozone-treated and Control Respirator Samples inoculated with Bacteria. **legend:** Top row: cultures from respirators inoculated with bacteria culture, exposed to 400 ppm ozone 80% humidity for two hours, and incubated for 24 hours. Bottom row: cultures from respirators inoculated with bacterial culture, exposed to ambient air 35% humidity for two hours, and incubated for 24 hours. Columns are labeled to identify respirator types tested. Tests were performed in duplicate for each respirator type. Serial dilutions were performed to enumerate the numbers of live bacteria.

**Table C1.**
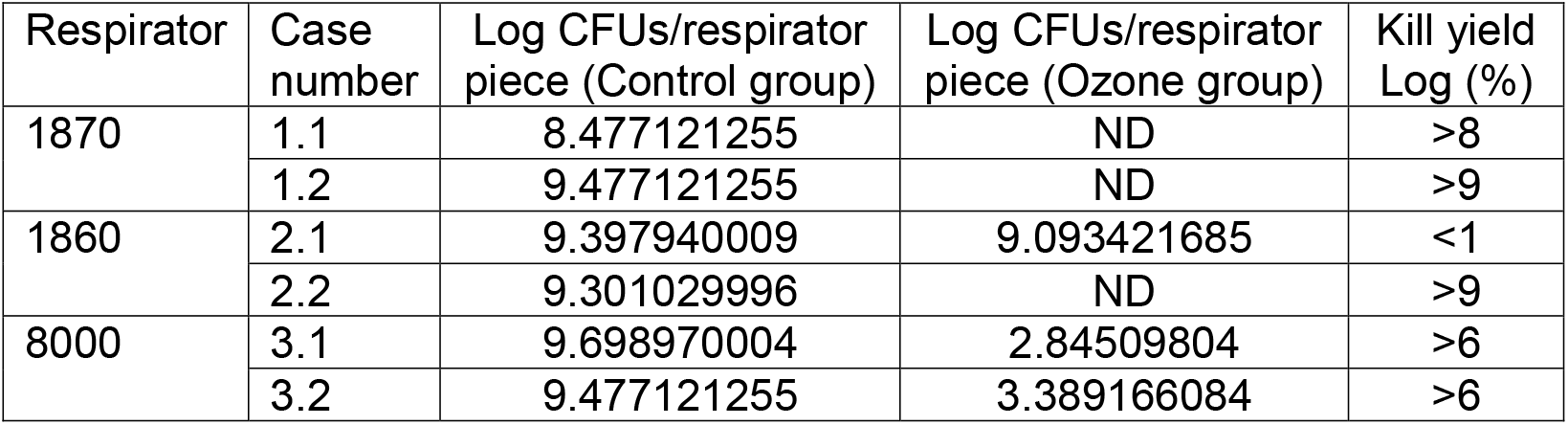
Log Kill Efficacy in Ozone-Exposed Respirator Pieces compared to Controls. **Key:** CFU = colony-forming units; ND = Not detected. Kill yield log is expressed as the base 10 logarithm of the reduction in organisms. Log 3 = 99.9%, log 6 = 99.9999%, etc.

### Supplemental Section D

Images from Filtration Efficiency Testing

Top: Image of N95 respirator after ten treatments with 450 ppm ozone for 2 hours at 75-90% humidity. There is little noticeable wear on the respirator after extended exposure to ozone. Bottom: Laboratory test photos of 1870 N95 respirators at CDC for quantitative fit testing using the TSI 8130A or mannikin testing.

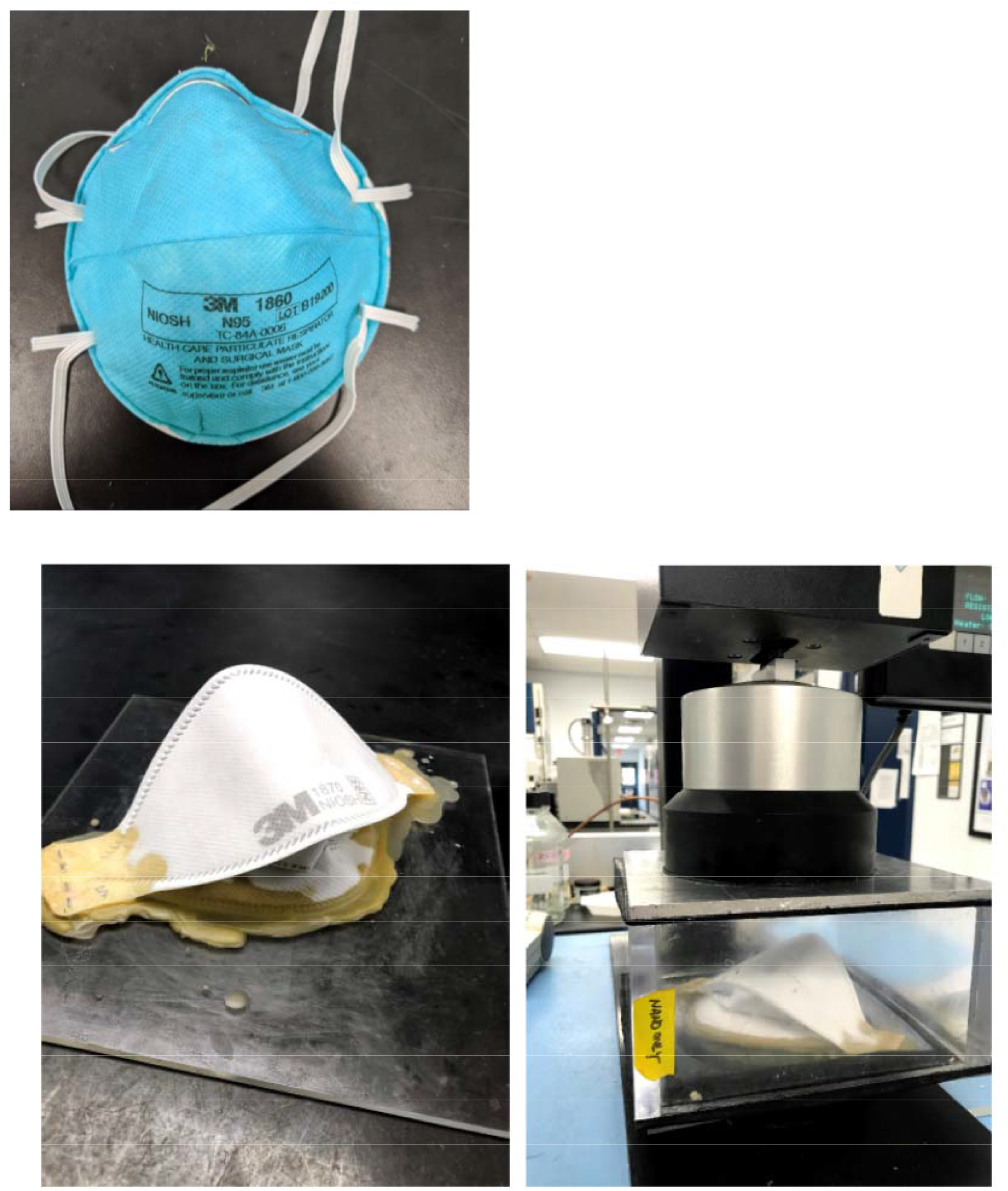
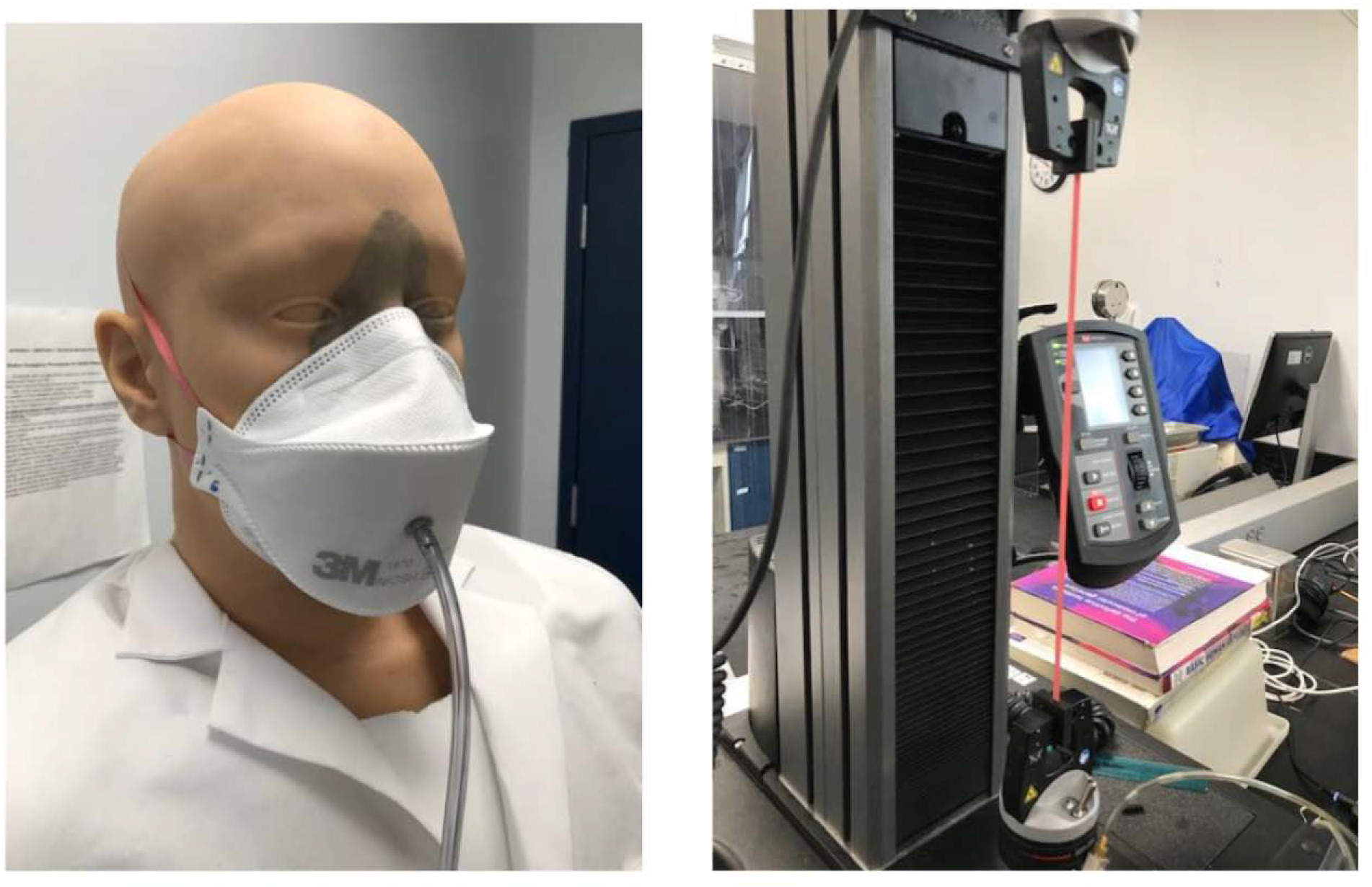

### Supplemental Section E

Picture of prototype ozone chamber inside fume hood for kill efficacy testing.

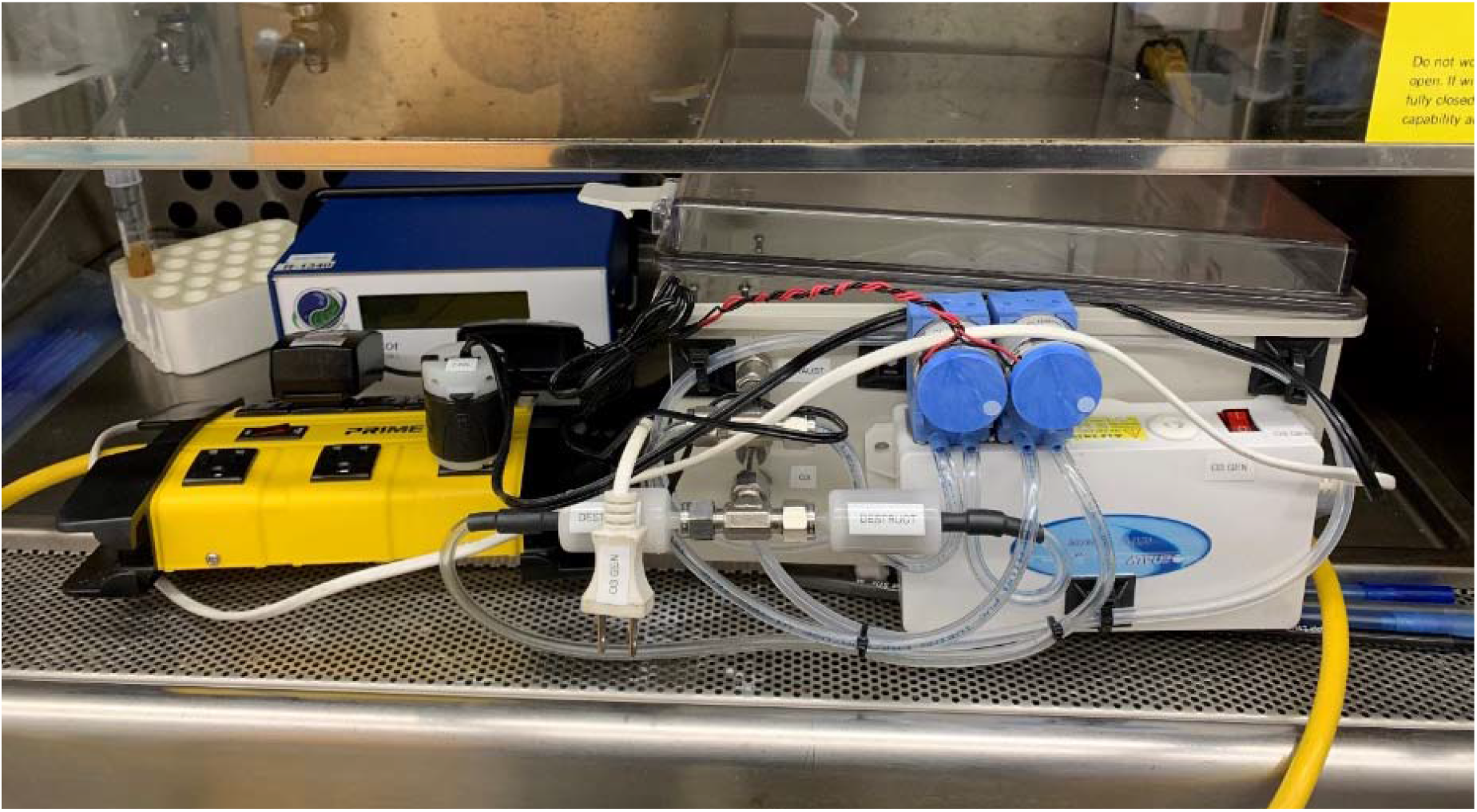

### Supplemental Section F

Ozone and Relative Humidity (RH) versus time for 1870 Respirators—5 treatment samples

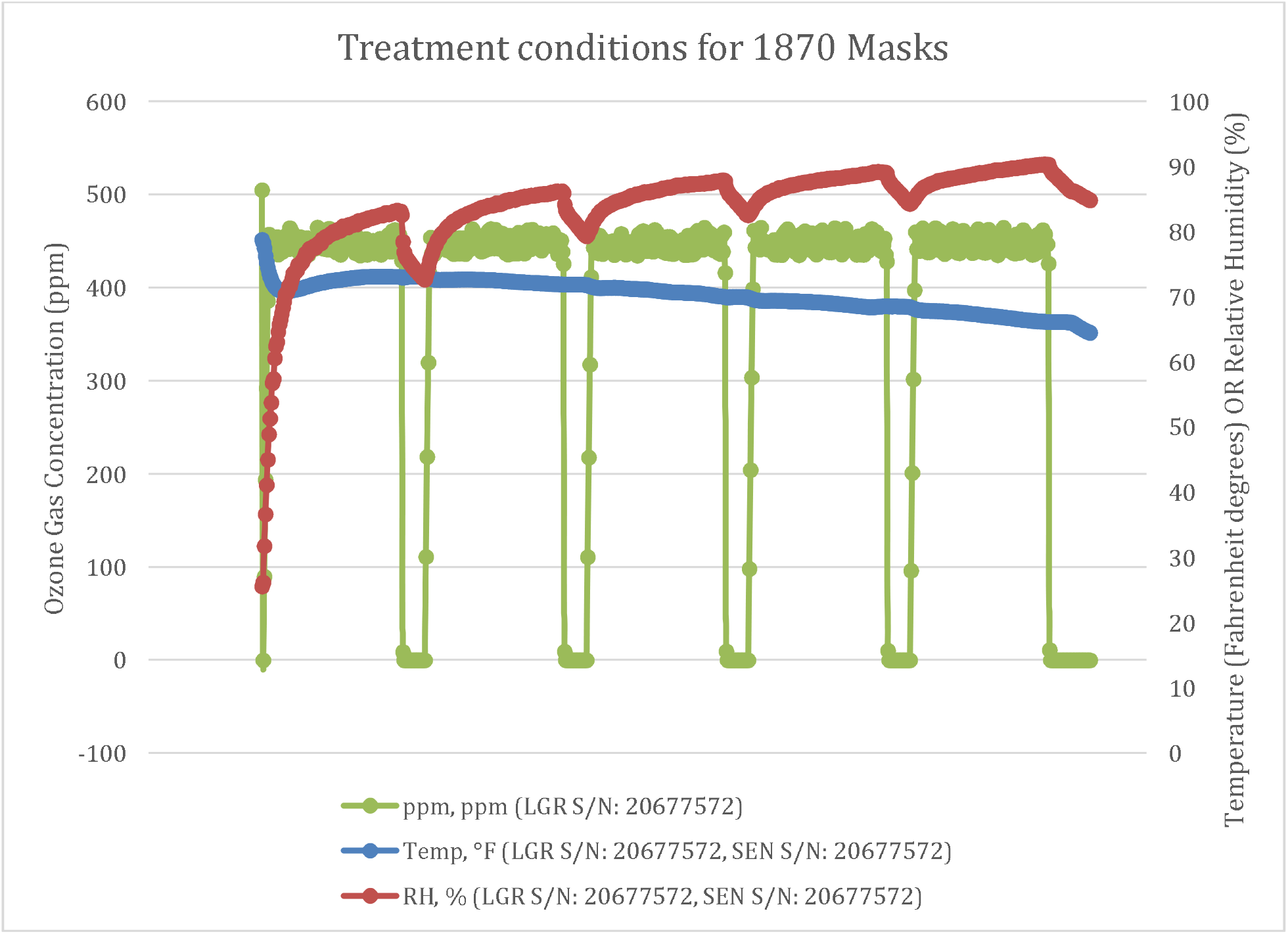

**Legend:** Traces showing the treatment conditions (ozone gas concentration, ambient temperature, and relative humidity (RH)) for the five 1870 masks that were cycled to 450 ppm for two hours each cycle at room temperature with humidity ranging from 75-90% during exposure periods. These masks were then sent to the CDC laboratory (Pittsburgh, PA) for filtration efficiency testing as detailed in the text.

## Notes

### Competing Interest Statement

The ozone chamber used for bacterial experiments was provided to the authors by Ozone Solutions (Hull, IA).
Sannel Patel is an employee of Ozone Solutions (Hull, IA).

### Author Declarations

No human subjects were involved in this study.

